# Health literacy, cognitive ability and self-reported diabetes in the English Longitudinal Study of Ageing

**DOI:** 10.1101/19003756

**Authors:** Chloe Fawns-Ritchie, Jackie Price, Ian J Deary

## Abstract

**Objective:** To examine the association of health literacy and cognitive ability with risk of diabetes.

Research Design and Methods: Participants were 8,669 English Longitudinal Study of Ageing participants (mean age 66.7 years, SD 9.7) who completed health literacy and cognitive ability tests at wave 2 (2004-2005), and who answered a self-reported question on whether a doctor had ever diagnosed them with diabetes. Logistic regression was used to examine the cross-sectional associations of health literacy and cognitive ability with diabetes status. In those without diabetes at wave 2, Cox regression was used to test the associations of health literacy and cognitive ability with risk of diabetes over a median of 9.5 years follow-up (n=6,961).

**Results:** Adequate (compared to limited) health literacy (OR 0.72, 95% CI 0.61-0.84) and higher cognitive ability (OR per 1 SD 0.73, CI 0.67-0.80) were both associated with lower odds of self-reported diabetes. Adequate health literacy (HR 0.64; CI 0.53-0.77) and higher cognitive ability (HR 0.77, CI 0.69-0.85) were also associated with lower risk of self-reporting diabetes during follow-up. When both health literacy and cognitive ability were added to the same model, these associations were slightly attenuated. Additional adjustment for health behaviours, education and social class attenuated associations further, and neither health literacy nor cognitive ability were significantly associated with diabetes.

**Conclusions:** Adequate health literacy and better cognitive ability were associated with reduced risk of diabetes. These associations were independent of each other, but not of other health- and socioeconomic-related variables.

---

Diabetes is a common chronic condition in older adulthood and is associated with substantial morbidity and mortality (1). Type 2 diabetes, the most common type of diabetes, is at least partly preventable (2). Understanding the characteristics of those most at risk of developing diabetes is important to appropriately target diabetes education and interventions. Known risk factors for developing diabetes include older age, deprivation, and obesity (1, 2).

Lower cognitive ability may be a risk factor for diabetes. Whereas one study (3) found that childhood cognitive ability did not predict diabetes in midlife, others have found that lower cognitive ability in early life was associated with higher risk of diabetes in adulthood (4, 5). In a sample of Scottish older adults who had their cognitive ability tested in childhood (4), a 1 SD advantage in cognitive ability was associated with 26% lower odds of reporting diabetes in older age. Individuals with higher cognitive ability might have the cognitive skills required to self-manage their health, take better care of themselves throughout life, and thus reduce the risk of developing diabetes (4, 6).

Health literacy, of which cognitive ability is thought to be a prerequisite (7), might also play a role in diabetes risk. Health literacy is the “capacity to obtain, process and understand basic health information and services needed to make basic health decisions” (8). In cross-sectional studies, rates of diabetes are higher in those with low health literacy (9, 10). In one study, participants with inadequate health literacy were 48% more likely to report having diabetes when compared to participants with adequate health literacy, even after adjusting for sociodemographic and health variables (10). Individuals with lower health literacy might not have the health-related skills required to obtain, understand and follow health advice, such as eating well and exercising, which might reduce the risk of diabetes (8).

In individuals with diabetes, higher health literacy has consistently been associated with greater diabetes knowledge (11-13). A very small association between higher health literacy and lower HbA1c levels in patients with diabetes has also been reported in a meta-analysis based on 26 studies (*r*=-0.048, *p*=0.027) (12). Whereas studies have investigated the association between health literacy and disease management in people with diabetes, little is known about whether health literacy is associated with risk of developing diabetes.

Health literacy and cognitive ability test scores are positively correlated (14, 15). In one study, rank-order correlations between a measure of general cognitive ability and three health literacy tests ranged from 0.37 to 0.50 (15). These correlations with cognitive ability tended to be higher than the correlations between the three health literacy tests themselves (*rho* = 0.28 to 0.46) (15). Researchers have sought to determine the role of cognitive ability in the association between health literacy and a range of health outcomes. Most (but not all (16)) studies have found that cognitive ability partly or entirely attenuates the association between health literacy and health (17-20). It is possible that any association between health literacy and diabetes may be attenuated when also measuring cognitive ability.

The aim of the current study was to better understand the associations of health literacy and cognitive ability with risk of diabetes. Using participants from the English Longitudinal Study of Ageing (ELSA), a cohort study designed to be representative of adults aged over 50 years living in England (21), the present study investigated whether health literacy and cognitive ability were independently associated with self-reported diabetes status. First, the cross-sectional associations between health literacy, cognitive ability, and self-reported diabetes were investigated. Second, individuals without diabetes at baseline were followed-up for up to 10 years to determine whether health literacy and cognitive ability were independently associated with subsequent risk of diabetes in mid-to-later life.

## RESEARCH DESIGN AND METHODS

### Participants

This study used data from core members of the ELSA study, a prospective cohort study of community-dwelling adults residing in England. The wave 1 (2002-2003) sample consisted of 11,391 participants who had previously participated in the Health Survey for England and who were living in a private household (21). ELSA participants have been followed up every two years.

The main interview consisted of participants answering questions on health, lifestyle and economic circumstances via a Computer Assisted Personal Interview (CAPI). Participants were asked to complete a self-completion questionnaire which assessed topics including diet and alcohol consumption. A nurse visit was carried out every second wave to assess physical measurements including height and weight, and blood and saliva samples were taken to measure biomarkers of disease. A detailed description of the sample design and the data collected in ELSA is reported elsewhere (21). The present study used data from waves 2 to 7. For the present study, baseline was considered to be Wave 2 (2004-2005; n=8,726), which was when the health literacy assessment was introduced.

Ethical approval for the English Longitudinal Study of Ageing was obtained from the National Research and Ethics Committee.

### Diabetes

Two measures of diabetes, collected during the CAPI, were used as outcome variables in this study.

#### Baseline diabetes status

Individuals who answered “yes” to “Has a doctor ever told you that you have diabetes?” at wave 2 were categorised as having diabetes. This question did not differentiate which type of diabetes the participant was diagnosed with. A previous study reported a high rate of agreement between self-reported diabetes and fasting blood glucose in a subsample of ELSA participants with data on both self-reported diabetes status and fasting blood glucose levels (22).

#### Incident diabetes

For incident diabetes, the analysis was restricted to participants who did not self-report diabetes at wave 2 and who had at least one wave of follow-up between waves 3 and 7. Participants who did not self-report diabetes at wave 2 and who subsequently answered “yes” to “Has a doctor ever told you that you have diabetes?” were categorised as having incident diabetes. As all participants were aged over 50 years at diagnosis, these cases are probably cases of type 2 diabetes.

#### Date of diabetes diagnosis

Individuals who self-reported diabetes were asked which month and year they were diagnosed. Date of diabetes diagnosis was used to calculate the time between wave 2 assessment and diabetes diagnosis.

### Health literacy

A brief 4-item health literacy test was administered during the CAPI at wave 2. This test assessed health-related reading comprehension skills which are thought to be required to successfully understand written health materials commonly encountered in healthcare. Participants were presented with a piece of paper containing a label for a packet of over-the-counter medication. Participants were asked four questions about the information on this label (e.g., “what is the maximum number of days you may take this medicine?”). The score was the number of correctly answered questions. As has been done in other studies (23, 24), performance was categorised as adequate (4/4 correct) or limited (<4 correct).

### Cognitive ability

Four tests administered during the wave 2 CAPI were used to create a measure of general cognitive ability. Immediate and delayed word recall were used to assess verbal declarative memory. In the immediate recall test, participants were read a list of 10 words and were asked to immediately recall as many of the words as possible. The score was the number of words recalled immediately. After a short delay, in which the words were not repeated, participants were asked to remember the 10 words again. The score was the number of words recalled after a delay. Executive function was assessed with a verbal fluency test. Participants were instructed to name as many animals as possible. The score was the number of animals named within the 60 second time limit. Letter cancellation was used to assess processing speed. Participants were presented with a piece of paper containing letters of the alphabet arranged in rows and columns. The task was to scan the piece of paper and score out all Ps and Ws. The score was the combined number of Ps and Ws scored out in 60 seconds.

Scores of 0 on animal fluency (n=48) and letter cancellation (n=3) were removed as this suggests participants did not complete the task or did not understand the task. Scores of 50 or more on animal fluency (n=4), and 60 or more on the letter cancellation (n=3) were removed as these scores were questionably high given the test time limits. Scores on the four cognitive ability tests were then entered into a principal component analysis (PCA). Only the first component had an eigenvalue >1, and the scree plot also indicated one component. Scores from the first principal component were saved and used as a measure of cognitive ability. The first component accounted for 57% of the variance in the scores on the four cognitive tests. The loadings were: Immediate word recall=0.83, delayed word recall=0.85, animal fluency=0.72, and letter cancellation=0.58. The resultant cognitive ability score was a z-score (mean 0.00, SD 1.00).

### Covariates

Age (in years), sex, BMI, health behaviours, number of cardiovascular comorbidities, and measures of socioeconomic status were used as covariates. Unless otherwise stated, all covariates were self-reported at the wave 2 CAPI. Participants aged over 90 years had their age set to 90 as there were so few of them. Participants were asked whether they smoked cigarettes nowadays and were categorised as current smokers or non-smokers. Participants were asked how often they took part in moderate and vigorous physical activity (more than once a week, once a week, one to three times a month, and hardly ever/never). Physical activity levels were categorised as vigorous activity at least once per week, moderate activity at least once per week, and physically inactive. Participants were asked about their frequency of alcohol consumption in the past 12 months in the self-completion questionnaire. This was categorised as never, rarely, at least once a month, at least once a week, and daily/almost daily. Height and weight, measured during the wave 2 nurse interview, were used to calculate BMI (kg/m^2^). Cardiovascular comorbidities were assessed by counting the number of self-reported cardiovascular conditions from hypertension, angina, heart attack, heart murmur, abnormal heart rhythm, stroke, and high cholesterol. Age that participants left full-time education was categorised as: age 14 or under, 15-16 years, 17-18 years, and age 19 or older. Social class was categorised using the National Statistics Socioeconomic Classification 3 categories (25): managerial and professional, intermediate, and routine and manual.

### Analysis

Independent t-tests were used to compare those with and without diabetes at wave 2 and those who did and did not develop diabetes at follow-up on normally-distributed continuous variables. Mann-Whitney U tests were used for non-normal continuous variables, and X^2^ tests were used for categorical variables. Spearman rank-order correlations were calculated between all predictor variables and co-variables.

Binary logistic regression was used to test the cross-sectional association of health literacy and cognitive ability with wave 2 diabetes status. Cox regression was used to investigate whether health literacy and cognitive ability test scores at wave 2 predicted risk of developing diabetes between waves 2 and 7. In the Cox regression analysis, time-to-event was taken as the difference, in days, between date of wave 2 CAPI and date of diabetes diagnosis for those who self-reported diabetes. For all other participants, time-to-event was the difference between date of wave 2 CAPI interview and the date of last CAPI interview. Month and year, but not day, were recorded for date of CAPI interview and date of diabetes diagnosis. To create a date variable (yyyy.mm.dd), the day was set to the middle of the month.

For the logistic regression and Cox regression, 6 models were run. Age and sex were entered into all models. Health literacy and cognitive ability were entered individually in models 1 and 2, respectively. Both health literacy and cognitive ability were added in Model 3 to determine whether the size of the health literacy-diabetes and cognitive ability-diabetes associations changed when concurrently entering both these variables in the model. To assess whether BMI and health behaviours accounted for these associations, BMI, smoking status, alcohol consumption, and physical activity were added in Model 4. Diabetes is a risk factor for cardiovascular disease (26). Associations between poorer cognitive ability and cardiovascular disease are also well established (27, 28). It is possible that any association between health literacy and cognitive ability with diabetes may be because of these associations with cardiovascular disease. To determine whether any association between health literacy and cognitive ability with diabetes was attenuated when adjusting for cardiovascular disease, number of cardiovascular comorbidities was additionally added in Model 5. Age of leaving full-time education and occupational social class were added in Model 6 to determine whether the association between health literacy, cognitive ability and diabetes was attenuated when accounting for these commonly-used indicators of socioeconomic status.

This study was interested in the associations of health literacy and cognitive ability with self-reported diabetes and the independence of these associations with respect to other health and socioeconomic-related variables. In the main text we report the ORs (95% CIs) and the HRs (95% CIs) for health literacy and cognitive ability only. The estimates for all covariates entered into the models are reported in the Supplementary materials.

## RESULTS

Of the 8,726 ELSA participants who completed the wave 2 assessment, 3 participants were removed who answered “don’t know” to whether a doctor had diagnosed them with diabetes. A further 54 were removed because these individuals selected that they had “diabetes or high blood sugar” from a Showcard listing cardiovascular conditions, but, when asked whether a doctor had ever told them they had diabetes, they answered “no”. Thus, the analytic sample consisted of 8,669 participants. Participant characteristics are reported in Table 1.

**Table 1.** Participant characteristics by diabetes status in the English Longitudinal Study of Ageing (ELSA)

|  | Baseline diabetes status reported at wave 2 of ELSA |  |  |  | Incident diabetes reported at follow-up* |  |  |  |
| --- | --- | --- | --- | --- | --- | --- | --- | --- |
|  | n | No diabetes<br>(n = 7961) | Diabetes<br>(n = 708) | p | n | No diabetes<br>(n = 6455) | Diabetes<br>(n = 506) | p |
| Age | 8669 | 66.46 (9.70) | 69.38 (9.16) | < .001 | 6961 | 66.02 (9.53) | 65.51 (8.59) | < .001 |
| Sex | 8669 |  |  | < .001 | 6961 |  |  | < .001 |
| Male |  | 3522 (44.2%) | 379 (53.5%) |  |  | 2791 (43.2%) | 262 (51.8%) |  |
| Female |  | 4439 (55.8%) | 329 (46.5%) |  |  | 3664 (56.8%) | 244 (48.2%) |  |
| Age left full-time education | 8468 |  |  | < .001 | 6809 |  |  | < .001 |
| <14 years |  | 1641 (21.1%) | 210 (30.6%) |  |  | 1222 (19.3%) | 107 (21.8%) |  |
| 15-16 years |  | 4085 (52.5%) | 349 (50.8%) |  |  | 3283 (52.0%) | 302 (61.6%) |  |
| 17-18 years |  | 1009 (13.0%) | 55 (8.0%) |  |  | 870 (13.8%) | 45 (9.2%) |  |
| 19+ years |  | 1046 (13.4%) | 73 (10.6%) |  |  | 944 (14.9%) | 36 (7.3%) |  |
| Social class | 8508 |  |  | < .001 | 6846 |  |  | < .001 |
| Managerial and professional |  | 2444 (31.2%) | 194 (28.4%) |  |  | 2067 (32.6%) | 133 (26.7%) |  |
| Intermediate |  | 1979 (25.3%) | 131 (19.2%) |  |  | 1662 (26.2%) | 104 (20.9%) |  |
| Routine and manual |  | 3403 (43.5%) | 357 (52.3%) |  |  | 2619 (41.3%) | 261 (52.4%) |  |
| Health literacy | 8293 |  |  | < .001 | 6736 |  |  | < .001 |
| Adequate |  | 5172 (67.7%) | 376 (57.8%) |  |  | 4351 (69.7%) | 300 (61.2%) |  |
| Limited |  | 2471 (32.3%) | 274 (42.2%) |  |  | 1895 (30.3%) | 190 (38.8%) |  |
| Cognitive ability | 8335 | 0.03 (1.00) | -0.36 (0.97) | < .001 | 6746 | 0.10 (0.98) | -0.04 (0.89) | < .001 |
| BMI | 7179 | 27.71 (4.79) | 30.45 (5.37) | < .001 | 5997 | 27.46 (4.64) | 31.21 (5.28) | < .001 |
| Current smoker | 8622 |  |  | 0.377 | 6929 |  |  | < .001 |
| Yes |  | 1216 (15.4%) | 99 (14.1%) |  |  | 934 (14.5%) | 105 (20.8%) |  |
| No |  | 6704 (84.6%) | 603 (85.9%) |  |  | 5490 (85.5%) | 400 (79.2%) |  |
| Alcohol | 7577 |  |  | < .001 | 6239 |  |  | < .001 |
| Never |  | 723 (10.3%) | 112 (19.3%) |  |  | 565 (9.7%) | 49 (11.2%) |  |
| Rarely |  | 1076 (15.4%) | 124 (21.3%) |  |  | 863 (14.9%) | 90 (20.6%) |  |
| At least once a month |  | 827 (11.8%) | 85 (14.6%) |  |  | 669 (11.5%) | 70 (16.1%) |  |
| At least once a week |  | 2662 (38.1%) | 171 (29.4%) |  |  | 2255 (38.9%) | 149 (34.2%) |  |
| Daily/almost daily |  | 1708 (24.4%) | 89 (15.3%) |  |  | 1451 (25.0%) | 78 (17.9%) |  |
| Physical activity | 8665 |  |  | < .001 | 6958 |  |  | < .001 |
| Vigorous activity |  | 2236 (28.1%) | 108 (15.2%) |  |  | 1938 (30.0%) | 116 (22.9%) |  |
| Moderate activity |  | 3888 (48.9%) | 305 (43.1%) |  |  | 3194 (49.5%) | 233 (46.0%) |  |
| Inactive |  | 1833 (23.0%) | 295 (41.7%) |  |  | 1320 (20.5%) | 157 (31.0%) |  |
| Number of cardiovascular comorbidities | 8669 | 0.67 (0.91) | 1.28 (1.13) | < .001 | 6961 | 0.64 (0.88) | 0.89 (1.04) | < .001 |

Scores are mean (SD) for continuous variables and count (percentage) for categorical variables. BMI, body mass index.
\*Incident diabetes reported at follow-up comparisons are based on a subsample of participants who did not self-report a diagnosis of diabetes at wave 2 and with at least one wave of follow-up.

### Baseline diabetes status

At baseline 708 (8.2%) of participants self-reported a doctor diagnosis of diabetes. Compared to those without diabetes, those with diabetes were more likely to have limited health literacy (42.2% versus 32.3%; *p*<.001) and have lower cognitive ability scores (diabetes mean -0.36, SD 0.97; no diabetes mean 0.03, SD 1.00; Cohen’s *d* 0.40; *p*<.001). Compared to participants without diabetes, participants with diabetes at wave 2 were older, more likely to be male, tended to leave full-time education at a younger age, be from a less professional social class, have a higher BMI, consume less alcohol, be inactive, and self-report more cardiovascular comorbidities. Rank-order correlations between the predictor variables and co-variables are reported in Table 2. Adequate health literacy was moderately correlated with higher scores on cognitive ability (*rho*=0.31, p<.001).

**Table 2.**
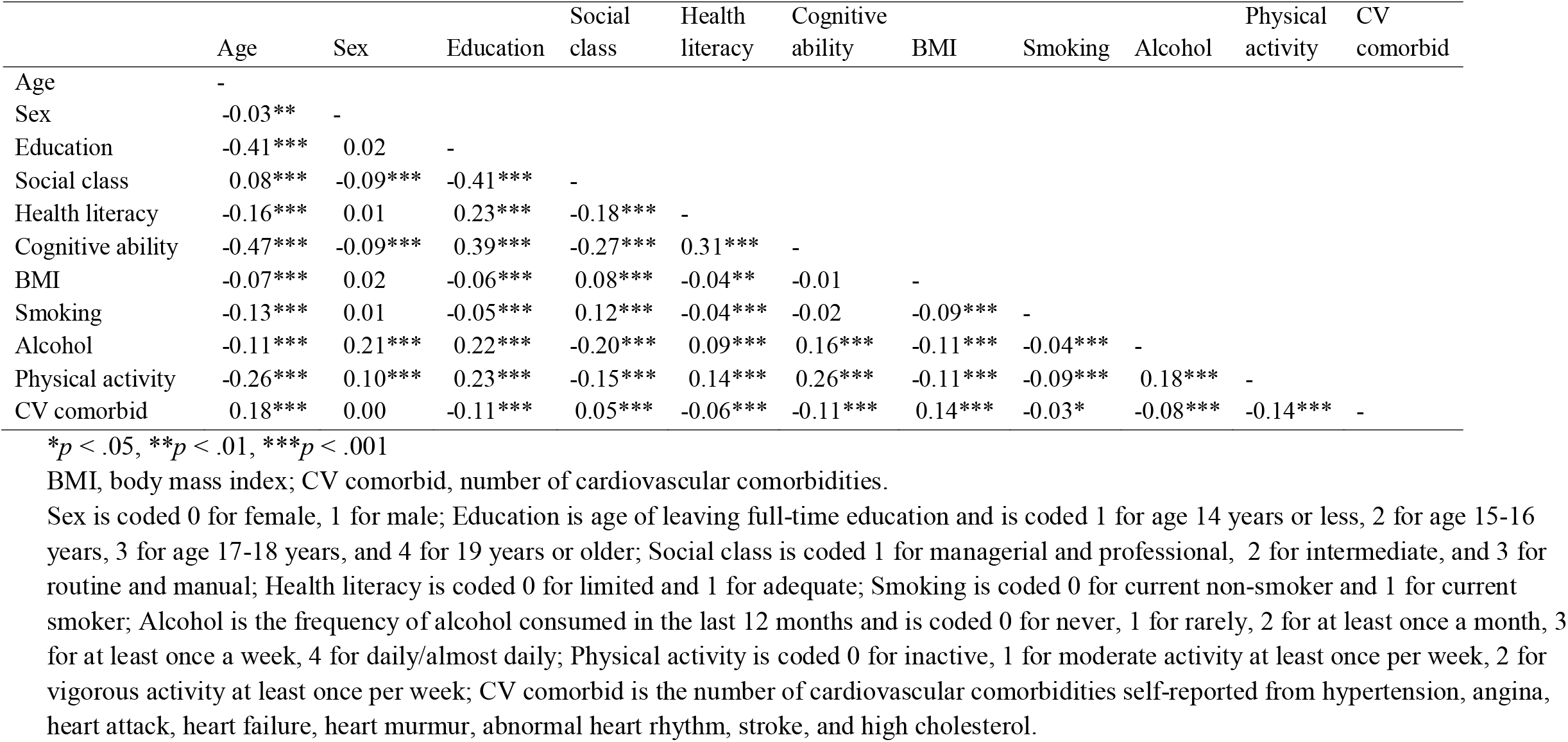
Spearman rank-order correlations between covariates (n=6,463 to 8,660)

The ORs and 95% CIs for the associations of health literacy and cognitive ability with self-reported diabetes at wave 2 are reported in Table 3 (full results are reported in Supplementary Table S1). A Box-Tidwell test found that the assumption of linearity of the logit was violated. Therefore an age-squared term was included in all models, and a squared term for number of cardiovascular comorbidities was included in models 5 and 6. Individuals with adequate health literacy were 28% less likely to self-report diabetes at wave 2 (Model 1; OR 0.72, 95% CI 0.61-0.84). A 1 SD higher cognitive ability was associated with a 27% lower odds of self-reporting diabetes (Model 2; OR 0.73, 95% CI 0.67-0.80). The association between health literacy and diabetes was attenuated by 36% (OR 0.82, 95% CI 0.69-0.98) and the association between cognitive ability and diabetes was attenuated by 19% (OR 0.78, 95% CI 0.70-0.86) when entering both health literacy and cognitive ability in Model 3. Both remained significantly associated with diabetes. The association between health literacy and cognitive ability with diabetes was attenuated and no longer significant when additionally adjusting for BMI and health behaviours in Model 4. Health literacy and cognitive ability remained non-significant after adjustment for cardiovascular comorbidities (Model 5), and for education and social class (Model 6).

**Table 3.** Odds ratios (95% CI) from logistic regression models of the association between health literacy and cognitive ability with whether participants self-reported a diagnosis of diabetes at wave 2 of the English Longitudinal Study of Ageing

|  | Model 1:<br>Health literacy | Model 2:<br>Cognitive ability | Model 3: Health<br>literacy and<br>cognitive ability | Model 4: + BMI<br>and health<br>behaviours | Model 5: + CV<br>comorbidities | Model 6: +<br>education and<br>social class |
| --- | --- | --- | --- | --- | --- | --- |
| Adequate health literacy | 0.72***<br>(0.61 to 0.84) | - | 0.82*<br>(0.69 to 0.98) | 0.97<br>(0.78 to 1.21) | 1.00<br>(0.81 to 1.26) | 0.98<br>(0.78 to 1.23) |
| Cognitive ability | - | 0.73***<br>(0.67 to 0.80) | 0.78***<br>(0.70 to 0.86) | 0.90<br>(0.80 to 1.02) | 0.90<br>(0.79 to 1.02) | 0.88<br>(0.77 to 1.00) |

\* $p < .05$ , \*\* $p < .01$ , \*\*\* $p < .001$
BMI, body mass index; CV, cardiovascular.
Models 1 (n=8,293), 2 (n=8,335), and 3 (n=8,185) adjusted for age, age-squared and sex. Model 4 (n=6,302) adjusted for age, age-squared, sex, body mass index, frequency of alcohol consumption in the past 12 months, and physical activity. Model 5 (n=6,302) adjusted for Model 4 covariates, number of cardiovascular comorbidities reported, and a squared term for number of cardiovascular comorbidities reported. Model 6 (n=6,086) adjusted for Model 5 covariates, age left full-time education, and occupational social class.

In the fully-adjusted model (Model 6; Supplementary Table S1) older age, being male, having a higher BMI, and reporting a higher number of cardiovascular comorbidities were associated with higher odds of having diabetes at wave 2. The association between number of cardiovascular comorbidities and diabetes became less strong as the number of comorbidities increased. Those who reported drinking alcohol at least once per month, rarely, or who never drank alcohol in the last 12 months were more likely to self-report diabetes when compared to those who reported drinking daily/almost daily. Compared to those who reported being physically inactive, those who took part in moderate or vigorous physical activity at least once per week were less likely to self-report diabetes.

### Risk of incident diabetes

Of the 7,961 participants who did not self-report diabetes at wave 2, 6,961 participants had at least one wave of follow-up between waves 3 and 7. They form the analytic sample for the association between health literacy, cognitive ability and risk of incident diabetes. A total of 506 (7.3%) participants reported a new diagnosis of diabetes between wave 3 and wave 7, whereas 6,455 (92.7%) participants did not. Median time to follow-up was 9.5 years. Mean time to censor for those with diabetes, who were censored at date of diabetes diagnosis, was 4.7 years (SD 3.1). Mean time to censor for those not diagnosed with diabetes, who were censored at date of last CAPI interview, was 7.8 years (SD 2.9). Characteristics for participants with and without diabetes at follow-up are shown in Table 1. Compared to participants who did not have incident diabetes, those who did were more likely to have limited health literacy (38.8% versus 30.3%, *p*<.001) and had lower cognitive ability scores (diabetes mean -0.04, SD 0.89; no diabetes mean 0.10, SD 0.98, Cohen’s *d* 0.15, *p*<.001) at wave 2. Compared to those who did not develop diabetes, participants who developed diabetes were older, more likely to be male, have left full-time education at a younger age, be from a less professional social class, smoke, consume less alcohol, be inactive, and to report more cardiovascular comorbidities at wave 2.

The HRs and 95% CIs for the association between health literacy, cognitive ability and risk of diabetes are reported in Table 4 (full results reported in Supplementary Table S2). Adequate health literacy at wave 2 was associated with a 36% lower risk of developing diabetes (Model 1; HR 0.64, 95% CI 0.53-0.77). A 1 SD higher cognitive ability score at wave 2 was associated with a 23% lower risk of developing diabetes (Model 2; HR 0.77, 95% CI 0.69-0.85). The association between health literacy and risk of diabetes was attenuated by 22% after adjustment for cognitive ability (Model 3; HR 0.72, 95% CI 0.59-0.87), and the association between cognitive ability and risk of diabetes was attenuated by 9% after adjusting for health literacy (HR 0.79, 95% CI 0.71-0.88). Both health literacy and cognitive ability remained significant predictors of diabetes risk. BMI and health behaviours were additionally added to the model in Model 4. The associations of health literacy (HR 0.79, 95% CI 0.64-0.99) and cognitive ability (HR 0.85, 95% CI 0.74-0.96) with diabetes risk were further attenuated but remained statistically significant. When additionally adjusting for number of cardiovascular comorbidities, the association between health literacy and cognitive ability with risk of diabetes remained almost unchanged (Model 5). After adjustment for age at leaving full-time education and social class, the associations of health literacy and cognitive ability with risk of diabetes were further attenuated and non-significant.

**Table 4.** Hazard ratios (95% CI) from Cox regression models of the association between health literacy and cognitive ability with risk of incident diabetes

|  | Model 1:<br>Health literacy | Model 2:<br>Cognitive ability | Model 3: Health<br>literacy and<br>cognitive ability | Model 4: + BMI<br>and health<br>behaviours | Model 5: + CV<br>comorbidities | Model 6: +<br>education and<br>social class |
| --- | --- | --- | --- | --- | --- | --- |
| Adequate health literacy | 0.64***<br>(0.53 to 0.77) | - | 0.72***<br>(0.59 to 0.87) | 0.79*<br>(0.64 to 0.99) | 0.80*<br>(0.64 to 0.99) | 0.85<br>(0.68 to 1.06) |
| Cognitive ability | - | 0.77***<br>(0.69 to 0.85) | 0.79***<br>(0.71 to 0.88) | 0.85*<br>(0.74 to 0.96) | 0.85*<br>(0.74 to 0.96) | 0.88<br>(0.77 to 1.01) |

\* $p < .05$ , \*\* $p < .01$ , \*\*\* $p < .001$
BMI, body mass index, CV, cardiovascular.
Models 1 (n=6,736; 490 diabetes events), 2 (n=6,746; 491 diabetes events), and 3 (n=6,654; 484 diabetes events) adjusted for age, and sex.
Model 4 (n=5,357; 377 diabetes events) adjusted for age, sex, body mass index, frequency of alcohol consumption in the past 12 months, and physical activity. Model 5 (n=5,357; 377 diabetes events) adjusted for Model 4 covariates, and number of cardiovascular comorbidities reported.
Model 6 (n=5,186; 360 diabetes events) adjusted for Model 5 covariates, age left full-time education, and occupational social class.

In the fully-adjusted model (Model 6; Supplementary Table S2) male participants, those with a higher BMI, current smokers, and those who reported consuming alcohol rarely (compared to those who reported consuming alcohol daily/almost daily) at wave 2 had an increased risk of diabetes. Participants who reported leaving education at age 19 years or older had a lower risk of diabetes when compared to those who left education at age 14 years or younger.

### Sensitivity analysis

There were some missing data. For the cross-sectional analyses, 70% of participants had complete data. For the longitudinal analyses, 75% of participants had complete data. All models were re-run using only participants with complete data on all variables entered into the models. These results are reported in Supplementary Tables S3 and S4. The pattern of associations were generally similar; however, the sizes of the associations tended to be slightly weaker compared to the full sample. For the cross-sectional analysis, health literacy was no longer significantly associated with diabetes status in Model 3 when adjusting for health literacy and cognitive ability (Supplementary Table S3). For the longitudinal analysis, when adjusting for BMI and health behaviours (Model 4; Supplementary Table S4), health literacy was no longer associated with risk of diabetes.

## CONCLUSIONS

Using a sample of middle-aged and older adults living in England, the present study found that adequate health literacy and better cognitive ability were associated with lower odds of reporting diabetes. The associations of health literacy and cognitive ability with diabetes status were attenuated and non-significant when additionally adjusting for BMI and health behaviours. Adequate health literacy and better cognitive ability, measured at wave 2, were associated with reduced risk of developing diabetes during a median of 9.5 years follow-up. Health literacy and cognitive ability predicted risk of diabetes both when examined individually and when examined concurrently. These associations were attenuated, though remained significant, when adjusting for BMI and health behaviours. When additionally adjusting for education and social class, the associations between health literacy and cognitive ability with diabetes risk were no longer significant. The results of the current study suggest that health literacy and cognitive ability have overlapping and unique associations with risk of diabetes. However, the relationship of health literacy and cognitive ability with diabetes is attenuated by health behaviours and education.

Previous cross-sectional studies have found that individuals with lower health literacy are more likely to report having diabetes (9, 10) and longitudinal studies have found that that lower cognitive ability earlier in life is associated with an increased risk of diabetes (4, 5). The present study is the first longitudinal study to examine whether health literacy is associated with risk of developing diabetes, and the first to examine whether cognitive ability and health literacy have independent associations with diabetes.

The association between health literacy and cognitive ability can at times be so strong that some have suggested that health literacy and cognitive ability are not unique constructs and, instead, that health literacy variance is mostly overlapping with cognitive ability (19; 29). If this were true, one would expect the association between health literacy and diabetes to be fully attenuated when adjusting for cognitive ability. This is not what was found in the current study. The association between health literacy and diabetes was only moderately attenuated (by 36% for baseline diabetes status and by 22% for diabetes risk) when adjusting for cognitive ability; moreover, both remained significant predictors of diabetes. The results suggest that only some of the association of health literacy and diabetes status with diabetes risk was accounted for by cognitive ability.

However, there is a necessary caveat to that possible conclusion. The cognitive ability measure created in the current study used four brief cognitive ability tests that assessed memory, executive function and processing speed, and did not include other important domains of cognitive function, such as reasoning, that are known to load highly on general cognitive ability (30). It is possible that some of the unique contribution of health literacy might be residual cognitive capability that was not picked up by the relatively brief measures of cognitive ability used (31). It is also not appropriate to compare the strength of the health literacy-diabetes and cognitive ability-diabetes associations due to the differences in measurement. Cognitive ability was assessed using a continuous measure created using scores on multiple cognitive ability tests, whereas health literacy scores were dichotomised into two groups (adequate and limited health literacy) based on performance on a brief, four-item test.

We found that the associations between health literacy and cognitive ability with cross-sectional diabetes status and with risk of diabetes were fully and partly attenuated, respectively, when adjusting for BMI and health behaviours. Better cognitive ability has been associated with health promoting behaviours such as following a healthy diet and taking part in regular exercise (3, 32-34). Whereas some studies have found associations between better health literacy and taking part in health promoting behaviours (35), others have not (36). Individuals with higher health literacy and cognitive ability might tend to be better equipped with the skills and abilities needed to take better care of themselves (6, 37) and to follow health advise including eating well and exercising, which, in turn, reduces the risk of developing diabetes (2).

Education attenuated and nullified the association between health literacy and cognitive ability with risk of diabetes. The association between better health literacy and cognitive ability with higher levels of education are well established (8, 38). Education may lead to better cognitive ability and health literacy, which in turn may lead to better health-related skills and lower rates of diabetes (19). Higher early life cognitive ability has been found to predict later educational attainment (38). Therefore, an alternative, but not mutually exclusive, explanation could be that higher cognitive ability may equip an individual with the skills needed to obtain higher educational qualifications. Higher educational attainment, in turn, may lead to better health (and lower risk of diabetes) by, for example, increasing health-related knowledge and decision-making skills (19). It is possible that including education in the models could be over-adjusting.

This study has a number of strengths and limitations. A key strength is that it examined the association of health literacy, cognitive ability and risk of incident diabetes longitudinally. Another strength is the relatively large sample size. One limitation is that only a subsample of participants had complete data. Those with missing data may be those with the lowest health literacy and cognitive ability scores. ELSA may also suffer from selective attrition such that those with increased risk of diabetes may be less likely to return for follow-up. Therefore, the results reported here may not generalise to those with the lowest health literacy and/or cognitive ability, and those with the highest risk of diabetes. Diabetes status was self-reported. However, there is a relatively high rate of agreement between self-reported diabetes and fasting blood glucose in ELSA (22). Only 1.7% of ELSA participants had undiagnosed diabetes, which is much lower than has been found in other cohort studies (22). The health literacy test used in the current study was a brief, four-item test which had limited variance (67% of participants scored the highest score). Although brief, this test was sensitive enough to predict diabetes risk in the current study, and it has previously been found to predict mortality (18).

This study found that both adequate health literacy and higher cognitive ability were independently associated with lower odds of self-reporting diabetes and with reduced risk of developing diabetes during a median of 9.5 years follow-up. These associations were attenuated by health behaviours and education. Individuals with poor health literacy and/or cognitive ability might tend to lack the health-related and cognitive skills and knowledge required to look after their health throughout life, which in turn, may increase the risk of diabetes. Future studies should investigate whether interventions designed to improve the knowledge and skills required to better self-manage health reduce the risk of developing diabetes in individuals with low health literacy and cognitive ability.

## Data Availability

The English Longitudinal Study of Ageing data can be downloaded from the UK Data Service (https://ukdataservice.ac.uk/).

## Acknowledgements

We thank our late colleague, John Starr, for his contribution to the conception and design of this study. The authors thank the participants of the ELSA study. We are grateful to the UK Data service for supplying the data.

This work was supported by the University of Edinburgh Centre for Cognitive Ageing and Cognitive Epidemiology, part of the cross council Lifelong Health and Wellbeing Initiative, funded by the Biotechnology and Biological Sciences Research Council (BBSRC), and Medical Research Council (MRC) (grant number MR/K026992/1).

## Author Contributions

CF-R discussed and planned the study, performed the statistical analysis, interpreted the data, and drafted the manuscript. JP discussed and planned the study, interpreted the data, and contributed to the manuscript. IJD discussed and planned the study, interpreted the data, and contributed to the manuscript. CF-R is the guarantor of this work and, as such, had full access to all the data in the study and takes responsibility for the integrity of the data and the accuracy of the data analysis.

## References

1. Diabetes UK: Diabetes: Facts and stats. 2014, p. 1–21

2. Hussain A, Claussen B, Ramachandran A, Williams R. Prevention of type 2 diabetes: a review. Diabetes Res Clin Pract 2007;76:317–326

3. Batty GD, Deary IJ, Macintyre S. Childhood IQ in relation to risk factors for premature mortality in middle-aged persons: the Aberdeen Children of the 1950s study. J Epidemiol Community Health 2007;61:241–247

4. Mõttus R, Luciano M, Starr JM, Deary IJ. Diabetes and life-long cognitive ability. J Psychosom Res 2013;75:275–278

5. Twig G, Gluzman I, Tirosh A, Gerstein HC, Yaniv G, Afek A, Derazne E, Tzur D, Karasik A, Gordon B, Fruchter E, Lubin G, Rudich A, Cukierman-Yaffe T. Cognitive function and the risk for diabetes among young men. Diabetes Care 2014;37:2982–2988

6. Gottfredson LS. Intelligence: is it the epidemiologists’ elusive “fundamental cause” of social class inequalities in health? J Pers Soc Psychol 2004;86:174–199

7. Paasche-Orlow MK, Parker RM, Gazmararian JA, Nielsen-Bohlman LT, Rudd RR. The prevalence of limited health literacy. J Gen Intern Med 2005;20:175–184

8. Nielsen-Bohlman L, Panzer AM, Kindig DA: Health Literacy: A Prescription to End Confusion. Washinton, DC, The National Academies Press, 2004

9. Adams RJ, Appleton SL, Hill CL, Dodd M, Findlay C, Wilson DH. Risks associated with low functional health literacy in an Australian population. Med J Aust 2009;191:530–534

10. Wolf MS, Gazmararian JA, Baker DW. Health literacy and functional health status among older adults. Arch Intern Med 2005;165:1946–1952

11. Caruso R, Magon A, Baroni I, Dellafiore F, Arrigoni C, Pittella F, Ausili D. Health literacy in type 2 diabetes patients: a systematic review of systematic reviews. Acta Diabetol 2018;55:1–12

12. Marciano L, Camerini AL, Schulz PJ. The Role of Health Literacy in Diabetes Knowledge, Self-Care, and Glycemic Control: a Meta-analysis. J Gen Intern Med 2019;

13. Al Sayah F, Majumdar SR, Williams B, Robertson S, Johnson JA. Health literacy and health outcomes in diabetes: a systematic review. J Gen Intern Med 2013;28:444–452

14. Bennett JS, Boyle PA, James BD, Bennett DA. Correlates of health and financial literacy in older adults without dementia. BMC Geriatr 2012;12:30

15. Murray C, Johnson W, Wolf MS, Deary IJ. The Association between Cognitive Ability across the Lifespan and Health Literacy in Old Age: The Lothian Birth Cohort 1936. Intelligence 2011;39:178–187

16. Lamar M, Wilson RS, Yu L, James BD, Stewart CC, Bennett DA, Boyle PA. Associations of literacy with diabetes indicators in older adults. J Epidemiol Community Health 2019;73:250–255

17. Baker DW, Wolf MS, Feinglass J, Thompson JA. Health literacy, cognitive abilities, and mortality among elderly persons. J Gen Intern Med 2008;23:723–726

18. Bostock S, Steptoe A. Association between low functional health literacy and mortality in older adults: longitudinal cohort study. BMJ 2012;344:e1602

19. Mõttus R, Johnson W, Murray C, Wolf MS, Starr JM, Deary IJ. Towards understanding the links between health literacy and physical health. Health Psychol 2014;33:164–173

20. Fawns-Ritchie C, Starr JM, Deary IJ. Role of cognitive ability in the association between functional health literacy and mortality in the Lothian Birth Cohort 1936: a prospective cohort study. BMJ Open 2018;8:e022502

21. Steptoe A, Breeze E, Banks J, Nazroo J. Cohort profile: the English longitudinal study of ageing. Int J Epidemiol 2013;42:1640–1648

22. Pierce MB, Zaninotto P, Steel N, Mindell J. Undiagnosed diabetes-data from the English longitudinal study of ageing. Diabet Med 2009;26:679–685

23. Kobayashi LC, Wardle J, von Wagner C. Limited health literacy is a barrier to colorectal cancer screening in England: evidence from the English Longitudinal Study of Ageing. Prev Med 2014;61:100–105

24. Gale CR, Deary IJ, Wardle J, Zaninotto P, Batty GD. Cognitive ability and personality as predictors of participation in a national colorectal cancer screening programme: the English Longitudinal Study of Ageing. J Epidemiol Community Health 2015;69:530

25. Rose D, Pevalin DJ, O’Reilly K: The National Statistics Socio-economic Classification: origins, development and use. Palgrave Macmillan Basingstoke, 2005

26. Sarwar N, Gao P, Seshasai SR, Gobin R, Kaptoge S, Di Angelantonio E, Ingelsson E, Lawlor DA, Selvin E, Stampfer M, Stehouwer CD, Lewington S, Pennells L, Thompson A, Sattar N, White IR, Ray KK, Danesh J. Diabetes mellitus, fasting blood glucose concentration, and risk of vascular disease: a collaborative meta-analysis of 102 prospective studies. Lancet 2010;375:2215–2222

27. Rostamian S, Mahinrad S, Stijnen T, Sabayan B, de Craen AJ. Cognitive impairment and risk of stroke: a systematic review and meta-analysis of prospective cohort studies. Stroke 2014;45:1342–1348

28. Hart CL, Taylor MD, Smith GD, Whalley LJ, Starr JM, Hole DJ, Wilson V, Deary IJ. Childhood IQ and cardiovascular disease in adulthood: prospective observational study linking the Scottish Mental Survey 1932 and the Midspan studies. Soc Sci Med (1982) 2004;59:2131–2138

29. Reeve CL, Basalik D. Is health literacy an example of construct proliferation? A conceptual and empirical evaluation of its redundancy with general cognitive ability. Intelligence 2014;44:93–102

30. Salthouse TA. Localizing age-related individual differences in a hierarchical structure. Intelligence 2004;32

31. Fawns-Ritchie C, Starr JM, Deary IJ. Health literacy, cognitive ability and smoking: a cross-sectional analysis of the English Longitudinal Study of Ageing. BMJ Open 2018;8:e023929

32. Batty GD, Deary IJ, Schoon I, Gale CR. Childhood mental ability in relation to food intake and physical activity in adulthood: the 1970 British Cohort Study. Pediatrics 2007;119:e38–45

33. Wraw C, Der G, Gale CR, Deary IJ. Intelligence in youth and health behaviours in middle age. Intelligence 2018;69:71–86

34. Batty GD, Deary IJ, Schoon I, Gale CR. Mental ability across childhood in relation to risk factors for premature mortality in adult life: the 1970 British Cohort Study. J Epidemiol Community Health 2007;61:997–1003

35. von Wagner C, Knight K, Steptoe A, Wardle J. Functional health literacy and health-promoting behaviour in a national sample of British adults. J Epidemiol Community Health 2007;61:1086

36. Wolf MS, Gazmararian JA, Baker DW. Health literacy and health risk behaviors among older adults. Am J Prev Med 2007;32:19–24

37. Deary IJ, Weiss A, Batty GD. Intelligence and Personality as Predictors of Illness and Death: How Researchers in Differential Psychology and Chronic Disease Epidemiology Are Collaborating to Understand and Address Health Inequalities. Psychol Sci Public Interest 2010;11:53–79

38. Deary IJ, Strand S, Smith P, Fernandes C. Intelligence and educational achievement. Intelligence 2007;35:13–21

